# Differences in COVID-19 Risk by Race and County-Level Social Determinants of Health Among Veterans

**DOI:** 10.1101/2021.10.20.21265149

**Authors:** Hoda S. Abdel Magid, Jacqueline M. Ferguson, Raymond V. Cleve, Amanda L. Purnell, Thomas F. Osborne

**Affiliations:** VA Palo Alto Healthcare System, US Department of Veterans Affairs, Palo Alto, CA, USA; Department of Epidemiology and Population Health, Stanford University, Stanford, CA, USA; Public Health Program, Santa Clara University, Santa Clara, CA, USA; Stanford Center for Population Health Sciences, Stanford University School of Medicine, Stanford, CA, USA; Veterans Affairs Central Office, Washington DC, USA; Department of Radiology, Stanford University, Stanford, CA, USA

**Keywords:** Veterans, COVID-19, Social Determinants of Health, County-Level, Race, Health Disparities

## Abstract

COVID-19 disparities by area-level social determinants of health (SDH) may be impacting U.S. Veterans. This retrospective analysis utilized COVID-19 data from the U.S. Department of Veterans Affairs (VA)’s EHR and geographically linked county-level data from 18 area-based socioeconomic measures. The risk of testing positive with Veterans’ county-level SDHs adjusting for demographics, comorbidities, and facility characteristics was calculated using generalized linear models. We found an exposure-response relationship whereby individual COVID-19 infection risk increased with each increasing quartile of adverse county-level SDH such as the percentage of residents in a county without a college degree, eligible for Medicaid, and living in crowded housing.

## Introduction

Disparities in COVID-19 infection and mortality vary across the US.^1-3^ These disparities, particularly among racial and ethnic minorities, may be driven by area-level social determinants of health (SDH) and structural resources.^4-6^ In this report, we combine electronic health record (EHR) data from the U.S. Department of Veterans Affairs (VA) with county-level characteristics to assess associations between area-level SDH and COVID-19 infection risk among Veterans with the goal of optimizing care and prevention strategies for our patients.

## Methods

We retrospectively examined records from Veterans actively enrolled in VA healthcare and who were tested for SARS-CoV-2 at VA between February 8, 2020 and December 28, 2020. Methods have been previously described in detail.^7^ In brief, we included demographic characteristics from the VA’s EHR database and used the Veteran’s home zip code to geographically link publicly available area-based SDH as it has been previously identified being critical for COVID-19 health equity in previous literature.^1,8-12^ A detailed table describing each county-level SDH, source, and original variable name from the source are provided in Supplementary **Table 1**. We categorized each area-based SDH into quartiles according to the positive case distribution in our analytic sample. We excluded Veterans missing county-level SDH and one VHA facility with fewer than 5 COVID-19 positive cases. The final analytic sample comprised 778,599 Veterans.

### Statistical Analysis

We used generalized linear models to report risk ratios and 95% confidence intervals for the risk of testing COVID-19 positive for key SDH. To examine effect modification by race between SDH and COVID-19 positivity risk, we stratified our analysis by race including White, Black, and Other Veterans (includes Asian, American Indian/Alaska Native, Native Hawaiian/Other Pacific Islander). All models were adjusted for individual demographics, facility characteristics, state, and other SDH characteristics that are important for health equity but not identified a-priori as primary SDH characteristics of interest.^1,8-12^ Model standard errors are clustered by VA facility. We conducted all statistical analyses using Stata Version 15 (StataCorp LLC). This quality assessment project received a Determination of Non-Research from Stanford Institutional Review Board as well as by VA determination.

## Results

As of December 28, 2020, among the 779,599 Veterans tested at VA, 77,692 (10%) tested positive for COVID-19. Compared with White Veterans, Black and Other Veterans on average lived in counties with higher percentages of non-US born residents, with higher percentage of non-White residents, individuals without a high school diploma, persons receiving food stamps/SNAP benefits; persons living in crowded housing, persons without broadband, persons living in multigenerational housing (i.e., households where grandparents have children who are under 18); and persons in deep poverty.

We found an exposure-response relationship with individual infection risk of COVID-19 increasing with each increasing quartile of adverse county-level SDH for the following SDH: percentage of residents in a county without a college degree, percentage eligible for Medicaid, and the percentage of residents living in crowded housing. (**Table 2**). The risk of testing positive for COVID-19 among Veterans living in counties with the top quartile of percentage of residents without a college degree compared to Veterans living in counties in the bottom quartile was 1.23 [95% confidence interval (CI): 1.10, 1.37]. Veterans living in the top quartile of counties with Medicaid eligibility were 1.17 [95% CI: 1.05, 1.37] times more likely to test positive for COVID-19 compared to Veterans living in the bottom quartile. Additionally, the relative risk of testing positive for COVID-19 among all Veterans living in the third quartile of crowded housing was 1.10 [95% CI: 1.04, 1.17] compared to the first quartile of persons in crowded housing. The association between county-level SDH and COVID-19 cases varied in race-stratified models. The relative risk for testing positive for COVID-19 among Black Veterans living in counties in the top versus bottom quartile of percentage of persons who are non-White was 1.16 (95% CI: 1.01, 1.33), however, among White Veterans the RR was attenuated (1.08 (95% CI: 0.95, 1.10)). Among Black Veterans living in counties in the top versus bottom quartile of percentage of households with multigenerational housing, the risk of testing positive for COVID-19 was 1.14 (95% CI: 1.04, 1.25), yet among White Veterans the RR was 1.01 (95% CI: 0.93, 1.10). Among Other Veterans, living in a county in the top versus bottom quartile of percentage of residents 25 years or older without 4+ years of college education was associated with a 31% (95% CI: 1.09-1.59) higher risk of testing positive for COVID-19 versus the lowest quartile. Comparing the top versus the bottom quartile, little to no differences were seen among the percentage of persons in deep poverty, percentage without a computer or broadband, and percentage non-US-born residents.

**Table 1.**
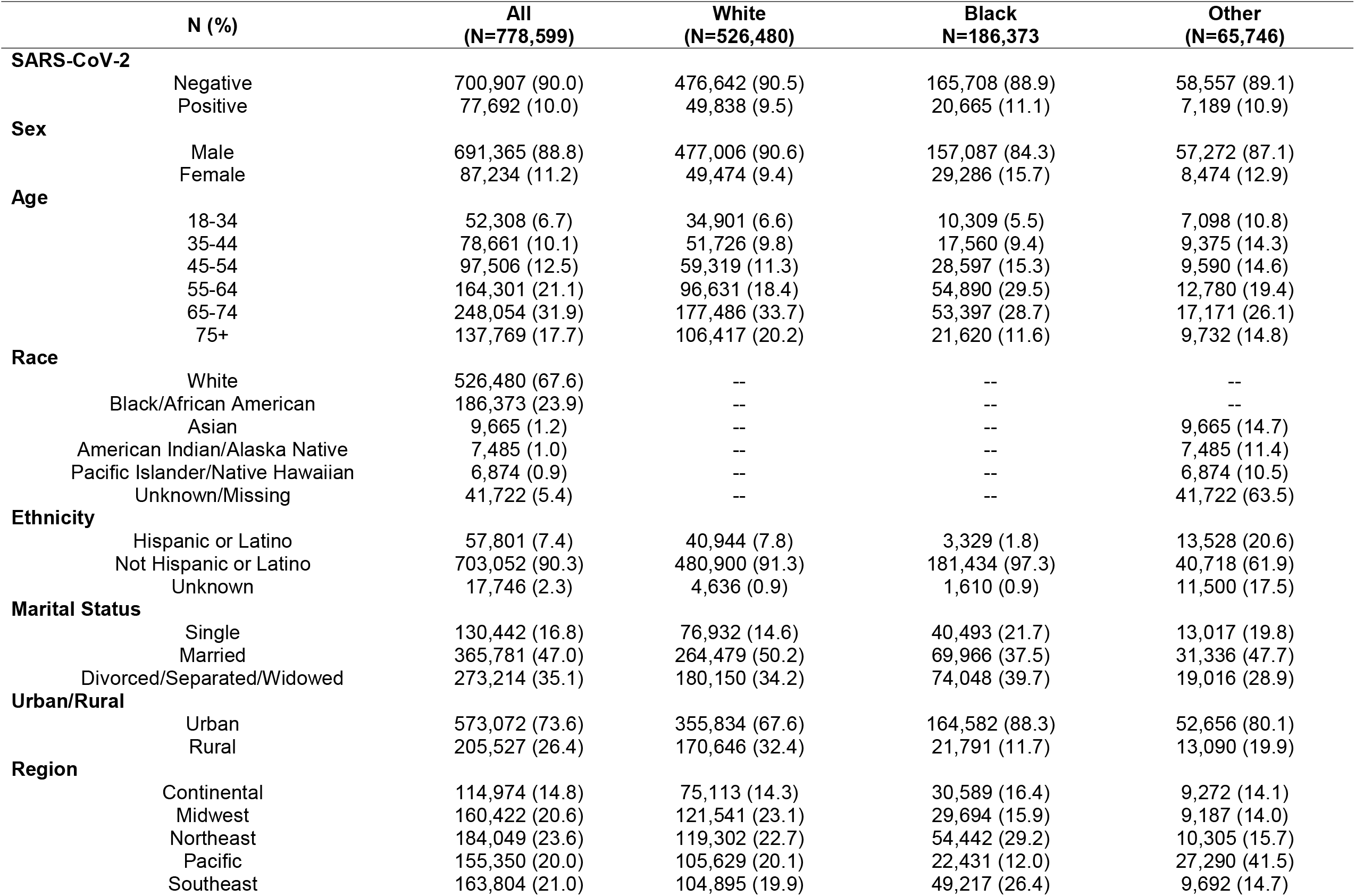
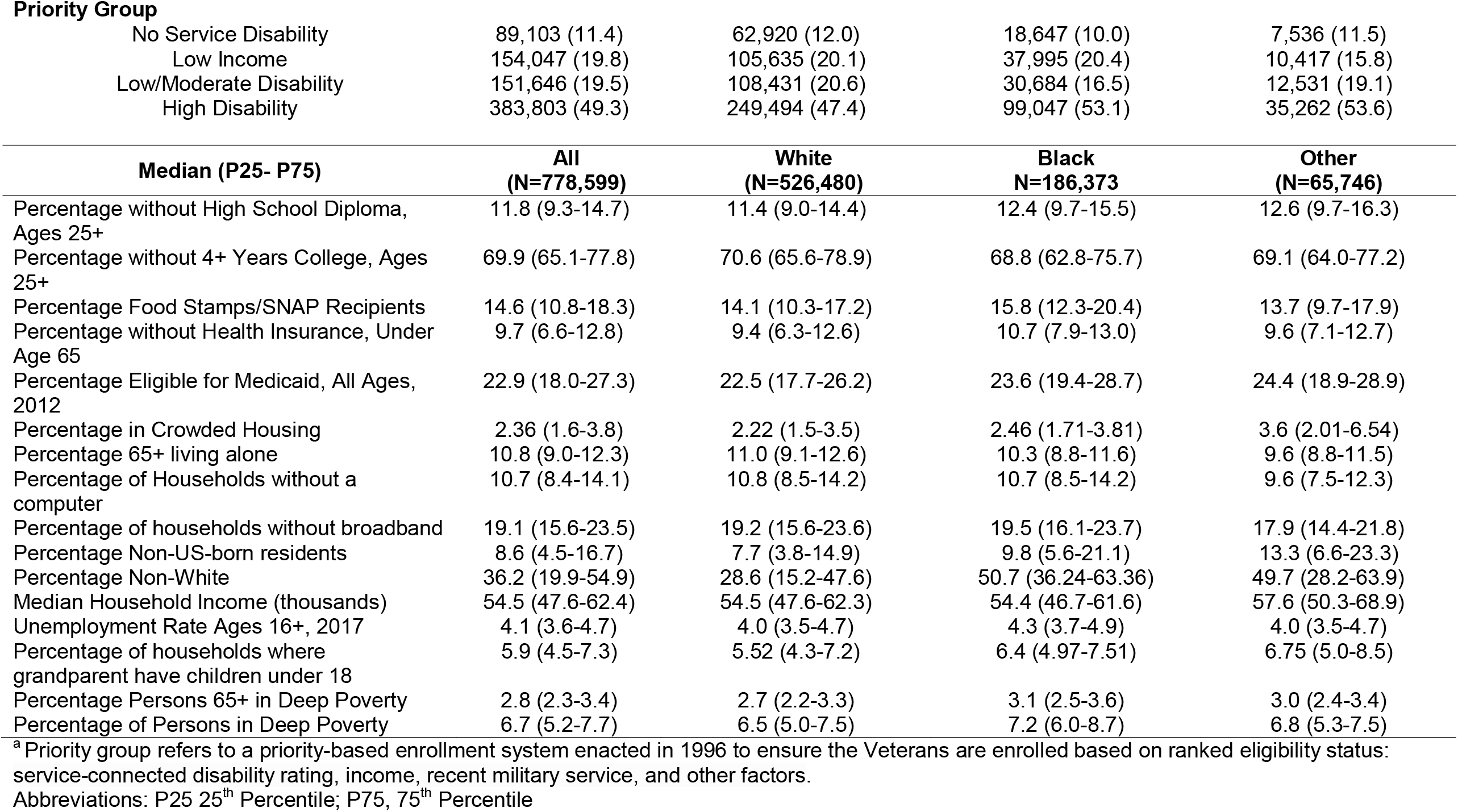
Individual and County-level Demographic and Social Determinants of Health Characteristics among U.S. Veterans enrolled in active care in the Veterans Health Administration (VHA), February 8 – December 28, 2020.

|  | N (%) | All<br>(N=778,599) | White<br>(N=526,480) | Black<br>N=186,373 | Other<br>(N=65,746) |
| --- | --- | --- | --- | --- | --- |
| **SARS-CoV-2** |  |  |  |  |  |
|  | Negative | 700,907 (90.0) | 476,642 (90.5) | 165,708 (88.9) | 58,557 (89.1) |
|  | Positive | 77,692 (10.0) | 49,838 (9.5) | 20,665 (11.1) | 7,189 (10.9) |
| **Sex** |  |  |  |  |  |
|  | Male | 691,365 (88.8) | 477,006 (90.6) | 157,087 (84.3) | 57,272 (87.1) |
|  | Female | 87,234 (11.2) | 49,474 (9.4) | 29,286 (15.7) | 8,474 (12.9) |
| **Age** |  |  |  |  |  |
|  | 18-34 | 52,308 (6.7) | 34,901 (6.6) | 10,309 (5.5) | 7,098 (10.8) |
|  | 35-44 | 78,661 (10.1) | 51,726 (9.8) | 17,560 (9.4) | 9,375 (14.3) |
|  | 45-54 | 97,506 (12.5) | 59,319 (11.3) | 28,597 (15.3) | 9,590 (14.6) |
|  | 55-64 | 164,301 (21.1) | 96,631 (18.4) | 54,890 (29.5) | 12,780 (19.4) |
|  | 65-74 | 248,054 (31.9) | 177,486 (33.7) | 53,397 (28.7) | 17,171 (26.1) |
|  | 75+ | 137,769 (17.7) | 106,417 (20.2) | 21,620 (11.6) | 9,732 (14.8) |
| **Race** |  |  |  |  |  |
|  | White | 526,480 (67.6) | -- | -- | -- |
|  | Black/African American | 186,373 (23.9) | -- | -- | -- |
|  | Asian | 9,665 (1.2) | -- | -- | 9,665 (14.7) |
|  | American Indian/Alaska Native | 7,485 (1.0) | -- | -- | 7,485 (11.4) |
|  | Pacific Islander/Native Hawaiian | 6,874 (0.9) | -- | -- | 6,874 (10.5) |
|  | Unknown/Missing | 41,722 (5.4) | -- | -- | 41,722 (63.5) |
| **Ethnicity** |  |  |  |  |  |
|  | Hispanic or Latino | 57,801 (7.4) | 40,944 (7.8) | 3,329 (1.8) | 13,528 (20.6) |
|  | Not Hispanic or Latino | 703,052 (90.3) | 480,900 (91.3) | 181,434 (97.3) | 40,718 (61.9) |
|  | Unknown | 17,746 (2.3) | 4,636 (0.9) | 1,610 (0.9) | 11,500 (17.5) |
| **Marital Status** |  |  |  |  |  |
|  | Single | 130,442 (16.8) | 76,932 (14.6) | 40,493 (21.7) | 13,017 (19.8) |
|  | Married | 365,781 (47.0) | 264,479 (50.2) | 69,966 (37.5) | 31,336 (47.7) |
|  | Divorced/Separated/Widowed | 273,214 (35.1) | 180,150 (34.2) | 74,048 (39.7) | 19,016 (28.9) |
| **Urban/Rural** |  |  |  |  |  |
|  | Urban | 573,072 (73.6) | 355,834 (67.6) | 164,582 (88.3) | 52,656 (80.1) |
|  | Rural | 205,527 (26.4) | 170,646 (32.4) | 21,791 (11.7) | 13,090 (19.9) |
| **Region** |  |  |  |  |  |
|  | Continental | 114,974 (14.8) | 75,113 (14.3) | 30,589 (16.4) | 9,272 (14.1) |
|  | Midwest | 160,422 (20.6) | 121,541 (23.1) | 29,694 (15.9) | 9,187 (14.0) |
|  | Northeast | 184,049 (23.6) | 119,302 (22.7) | 54,442 (29.2) | 10,305 (15.7) |
|  | Pacific | 155,350 (20.0) | 105,629 (20.1) | 22,431 (12.0) | 27,290 (41.5) |
|  | Southeast | 163,804 (21.0) | 104,895 (19.9) | 49,217 (26.4) | 9,692 (14.7) |

**Priority Group**
|  |  |  |  |  |
| --- | --- | --- | --- | --- |
| No Service Disability | 89,103 (11.4) | 62,920 (12.0) | 18,647 (10.0) | 7,536 (11.5) |
| Low Income | 154,047 (19.8) | 105,635 (20.1) | 37,995 (20.4) | 10,417 (15.8) |
| Low/Moderate Disability | 151,646 (19.5) | 108,431 (20.6) | 30,684 (16.5) | 12,531 (19.1) |
| High Disability | 383,803 (49.3) | 249,494 (47.4) | 99,047 (53.1) | 35,262 (53.6) |

| **Median (P25- P75)** | **All**<br>**(N=778,599)** | **White**<br>**(N=526,480)** | **Black**<br>**N=186,373** | **Other**<br>**(N=65,746)** |
| --- | --- | --- | --- | --- |
| Percentage without High School Diploma, Ages 25+ | 11.8 (9.3-14.7) | 11.4 (9.0-14.4) | 12.4 (9.7-15.5) | 12.6 (9.7-16.3) |
| Percentage without 4+ Years College, Ages 25+ | 69.9 (65.1-77.8) | 70.6 (65.6-78.9) | 68.8 (62.8-75.7) | 69.1 (64.0-77.2) |
| Percentage Food Stamps/SNAP Recipients | 14.6 (10.8-18.3) | 14.1 (10.3-17.2) | 15.8 (12.3-20.4) | 13.7 (9.7-17.9) |
| Percentage without Health Insurance, Under Age 65 | 9.7 (6.6-12.8) | 9.4 (6.3-12.6) | 10.7 (7.9-13.0) | 9.6 (7.1-12.7) |
| Percentage Eligible for Medicaid, All Ages, 2012 | 22.9 (18.0-27.3) | 22.5 (17.7-26.2) | 23.6 (19.4-28.7) | 24.4 (18.9-28.9) |
| Percentage in Crowded Housing | 2.36 (1.6-3.8) | 2.22 (1.5-3.5) | 2.46 (1.71-3.81) | 3.6 (2.01-6.54) |
| Percentage 65+ living alone | 10.8 (9.0-12.3) | 11.0 (9.1-12.6) | 10.3 (8.8-11.6) | 9.6 (8.8-11.5) |
| Percentage of Households without a computer | 10.7 (8.4-14.1) | 10.8 (8.5-14.2) | 10.7 (8.5-14.2) | 9.6 (7.5-12.3) |
| Percentage of households without broadband | 19.1 (15.6-23.5) | 19.2 (15.6-23.6) | 19.5 (16.1-23.7) | 17.9 (14.4-21.8) |
| Percentage Non-US-born residents | 8.6 (4.5-16.7) | 7.7 (3.8-14.9) | 9.8 (5.6-21.1) | 13.3 (6.6-23.3) |
| Percentage Non-White | 36.2 (19.9-54.9) | 28.6 (15.2-47.6) | 50.7 (36.24-63.36) | 49.7 (28.2-63.9) |
| Median Household Income (thousands) | 54.5 (47.6-62.4) | 54.5 (47.6-62.3) | 54.4 (46.7-61.6) | 57.6 (50.3-68.9) |
| Unemployment Rate Ages 16+, 2017 | 4.1 (3.6-4.7) | 4.0 (3.5-4.7) | 4.3 (3.7-4.9) | 4.0 (3.5-4.7) |
| Percentage of households where grandparent have children under 18 | 5.9 (4.5-7.3) | 5.52 (4.3-7.2) | 6.4 (4.97-7.51) | 6.75 (5.0-8.5) |
| Percentage Persons 65+ in Deep Poverty | 2.8 (2.3-3.4) | 2.7 (2.2-3.3) | 3.1 (2.5-3.6) | 3.0 (2.4-3.4) |
| Percentage of Persons in Deep Poverty | 6.7 (5.2-7.7) | 6.5 (5.0-7.5) | 7.2 (6.0-8.7) | 6.8 (5.3-7.5) |

<sup>a</sup> Priority group refers to a priority-based enrollment system enacted in 1996 to ensure the Veterans are enrolled based on ranked eligibility status: service-connected disability rating, income, recent military service, and other factors.
Abbreviations: P25 25<sup>th</sup> Percentile; P75, 75<sup>th</sup> Percentile

**Table 2.**
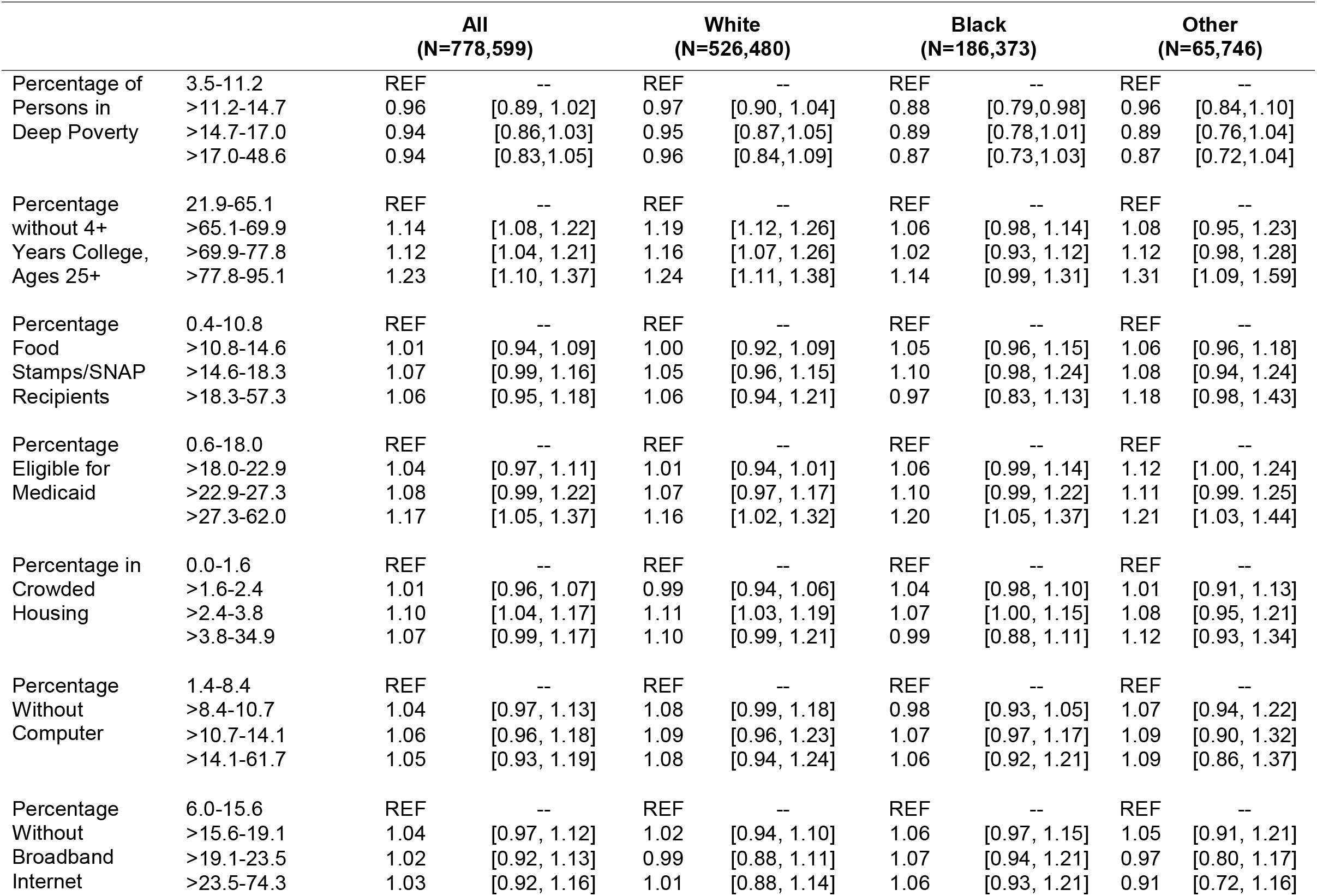
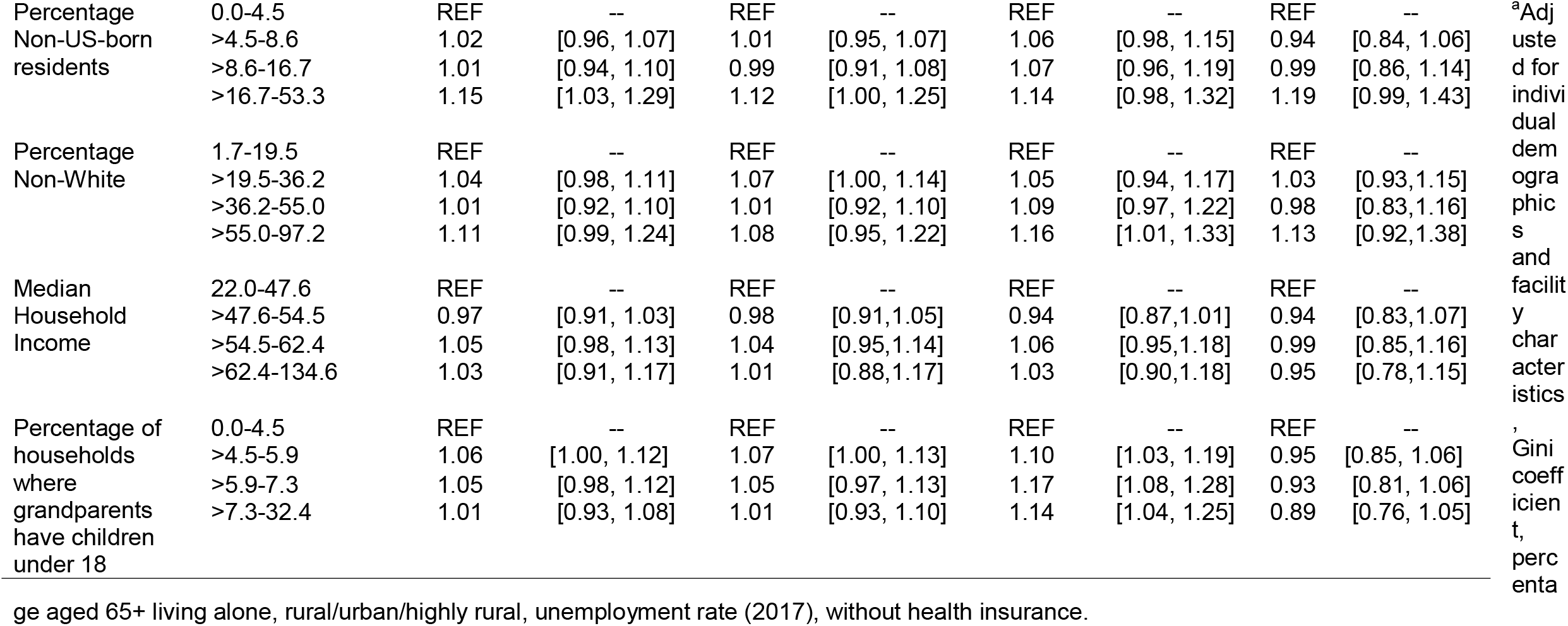
Adjusted risk ratios (95 CI) for receiving a positive COVID-19 test result among Veterans Enrolled in Active Care at the Veterans Health Administration (VHA) who obtained a COVID-19 test, February 8–December 28, 2020.^a^

|  |  | **All**<br>**(N=778,599)** |  | **White**<br>**(N=526,480)** |  | **Black**<br>**(N=186,373)** |  | **Other**<br>**(N=65,746)** |  |
| --- | --- | --- | --- | --- | --- | --- | --- | --- | --- |
| Percentage of<br>Persons in<br>Deep Poverty | 3.5-11.2 | REF | -- | REF | -- | REF | -- | REF | -- |
|  | >11.2-14.7 | 0.96 | [0.89, 1.02] | 0.97 | [0.90, 1.04] | 0.88 | [0.79,0.98] | 0.96 | [0.84,1.10] |
|  | >14.7-17.0 | 0.94 | [0.86,1.03] | 0.95 | [0.87,1.05] | 0.89 | [0.78,1.01] | 0.89 | [0.76,1.04] |
|  | >17.0-48.6 | 0.94 | [0.83,1.05] | 0.96 | [0.84,1.09] | 0.87 | [0.73,1.03] | 0.87 | [0.72,1.04] |
| Percentage<br>without 4+<br>Years College,<br>Ages 25+ | 21.9-65.1 | REF | -- | REF | -- | REF | -- | REF | -- |
|  | >65.1-69.9 | 1.14 | [1.08, 1.22] | 1.19 | [1.12, 1.26] | 1.06 | [0.98, 1.14] | 1.08 | [0.95, 1.23] |
|  | >69.9-77.8 | 1.12 | [1.04, 1.21] | 1.16 | [1.07, 1.26] | 1.02 | [0.93, 1.12] | 1.12 | [0.98, 1.28] |
|  | >77.8-95.1 | 1.23 | [1.10, 1.37] | 1.24 | [1.11, 1.38] | 1.14 | [0.99, 1.31] | 1.31 | [1.09, 1.59] |
| Percentage<br>Food<br>Stamps/SNAP<br>Recipients | 0.4-10.8 | REF | -- | REF | -- | REF | -- | REF | -- |
|  | >10.8-14.6 | 1.01 | [0.94, 1.09] | 1.00 | [0.92, 1.09] | 1.05 | [0.96, 1.15] | 1.06 | [0.96, 1.18] |
|  | >14.6-18.3 | 1.07 | [0.99, 1.16] | 1.05 | [0.96, 1.15] | 1.10 | [0.98, 1.24] | 1.08 | [0.94, 1.24] |
|  | >18.3-57.3 | 1.06 | [0.95, 1.18] | 1.06 | [0.94, 1.21] | 0.97 | [0.83, 1.13] | 1.18 | [0.98, 1.43] |
| Percentage<br>Eligible for<br>Medicaid | 0.6-18.0 | REF | -- | REF | -- | REF | -- | REF | -- |
|  | >18.0-22.9 | 1.04 | [0.97, 1.11] | 1.01 | [0.94, 1.01] | 1.06 | [0.99, 1.14] | 1.12 | [1.00, 1.24] |
|  | >22.9-27.3 | 1.08 | [0.99, 1.22] | 1.07 | [0.97, 1.17] | 1.10 | [0.99, 1.22] | 1.11 | [0.99, 1.25] |
|  | >27.3-62.0 | 1.17 | [1.05, 1.37] | 1.16 | [1.02, 1.32] | 1.20 | [1.05, 1.37] | 1.21 | [1.03, 1.44] |
| Percentage in<br>Crowded<br>Housing | 0.0-1.6 | REF | -- | REF | -- | REF | -- | REF | -- |
|  | >1.6-2.4 | 1.01 | [0.96, 1.07] | 0.99 | [0.94, 1.06] | 1.04 | [0.98, 1.10] | 1.01 | [0.91, 1.13] |
|  | >2.4-3.8 | 1.10 | [1.04, 1.17] | 1.11 | [1.03, 1.19] | 1.07 | [1.00, 1.15] | 1.08 | [0.95, 1.21] |
|  | >3.8-34.9 | 1.07 | [0.99, 1.17] | 1.10 | [0.99, 1.21] | 0.99 | [0.88, 1.11] | 1.12 | [0.93, 1.34] |
| Percentage<br>Without<br>Computer | 1.4-8.4 | REF | -- | REF | -- | REF | -- | REF | -- |
|  | >8.4-10.7 | 1.04 | [0.97, 1.13] | 1.08 | [0.99, 1.18] | 0.98 | [0.93, 1.05] | 1.07 | [0.94, 1.22] |
|  | >10.7-14.1 | 1.06 | [0.96, 1.18] | 1.09 | [0.96, 1.23] | 1.07 | [0.97, 1.17] | 1.09 | [0.90, 1.32] |
|  | >14.1-61.7 | 1.05 | [0.93, 1.19] | 1.08 | [0.94, 1.24] | 1.06 | [0.92, 1.21] | 1.09 | [0.86, 1.37] |
| Percentage<br>Without<br>Broadband<br>Internet | 6.0-15.6 | REF | -- | REF | -- | REF | -- | REF | -- |
|  | >15.6-19.1 | 1.04 | [0.97, 1.12] | 1.02 | [0.94, 1.10] | 1.06 | [0.97, 1.15] | 1.05 | [0.91, 1.21] |
|  | >19.1-23.5 | 1.02 | [0.92, 1.13] | 0.99 | [0.88, 1.11] | 1.07 | [0.94, 1.21] | 0.97 | [0.80, 1.17] |
|  | >23.5-74.3 | 1.03 | [0.92, 1.16] | 1.01 | [0.88, 1.14] | 1.06 | [0.93, 1.21] | 0.91 | [0.72, 1.16] |

|  |  |  |  |  |  |  |  |  |  |  |
| --- | --- | --- | --- | --- | --- | --- | --- | --- | --- | --- |
| Percentage Non-US-born residents | 0.0-4.5<br>>4.5-8.6<br>>8.6-16.7<br>>16.7-53.3 | REF<br>1.02<br>1.01<br>1.15 | --<br>[0.96, 1.07]<br>[0.94, 1.10]<br>[1.03, 1.29] | REF<br>1.01<br>0.99<br>1.12 | --<br>[0.95, 1.07]<br>[0.91, 1.08]<br>[1.00, 1.25] | REF<br>1.06<br>1.07<br>1.14 | --<br>[0.98, 1.15]<br>[0.96, 1.19]<br>[0.98, 1.32] | REF<br>0.94<br>0.99<br>1.19 | --<br>[0.84, 1.06]<br>[0.86, 1.14]<br>[0.99, 1.43] | <sup>a</sup>Adjusted for individual demographics and facility characteristics, Gini coefficient, percentage |
| Percentage Non-White | 1.7-19.5<br>>19.5-36.2<br>>36.2-55.0<br>>55.0-97.2 | REF<br>1.04<br>1.01<br>1.11 | --<br>[0.98, 1.11]<br>[0.92, 1.10]<br>[0.99, 1.24] | REF<br>1.07<br>1.01<br>1.08 | --<br>[1.00, 1.14]<br>[0.92, 1.10]<br>[0.95, 1.22] | REF<br>1.05<br>1.09<br>1.16 | --<br>[0.94, 1.17]<br>[0.97, 1.22]<br>[1.01, 1.33] | REF<br>1.03<br>0.98<br>1.13 | --<br>[0.93,1.15]<br>[0.83,1.16]<br>[0.92,1.38] |  |
| Median Household Income | 22.0-47.6<br>>47.6-54.5<br>>54.5-62.4<br>>62.4-134.6 | REF<br>0.97<br>1.05<br>1.03 | --<br>[0.91, 1.03]<br>[0.98, 1.13]<br>[0.91, 1.17] | REF<br>0.98<br>1.04<br>1.01 | --<br>[0.91,1.05]<br>[0.95,1.14]<br>[0.88,1.17] | REF<br>0.94<br>1.06<br>1.03 | --<br>[0.87,1.01]<br>[0.95,1.18]<br>[0.90,1.18] | REF<br>0.94<br>0.99<br>0.95 | --<br>[0.83,1.07]<br>[0.85,1.16]<br>[0.78,1.15] |  |
| Percentage of households where grandparents have children under 18 | 0.0-4.5<br>>4.5-5.9<br>>5.9-7.3<br>>7.3-32.4 | REF<br>1.06<br>1.05<br>1.01 | --<br>[1.00, 1.12]<br>[0.98, 1.12]<br>[0.93, 1.08] | REF<br>1.07<br>1.05<br>1.01 | --<br>[1.00, 1.13]<br>[0.97, 1.13]<br>[0.93, 1.10] | REF<br>1.10<br>1.17<br>1.14 | --<br>[1.03, 1.19]<br>[1.08, 1.28]<br>[1.04, 1.25] | REF<br>0.95<br>0.93<br>0.89 | --<br>[0.85, 1.06]<br>[0.81, 1.06]<br>[0.76, 1.05] |  |

ge aged 65+ living alone, rural/urban/highly rural, unemployment rate (2017), without health insurance.

## Discussion

Our results show that Veterans living in areas with lower education levels, higher Medicaid eligibility, crowded housing, non-White residents, and multigenerational housing are experiencing higher risks of COVID-19 infection, a trend which has been noted in other evaluations.^1,3,10,13^ Notably our assessment revealed important associations for our Veterans, such as percentage of residents who are non-White, multigenerational housing, and percentage of residents without a college degree varied in race-stratified models, strengthening for Black and Other Veterans, compared to White Veterans which provides important insights for our targeted interventions.

A strength of our work is that our findings also demonstrate the association between distinct county-level SDH and COVID-19 cases which was possible due to the large cohort size from a nationwide database from the largest integrated healthcare system in the United States. Moreover, our assessment was designed to provide a more precise evaluation to direct targeted enhancement for of our patients which was also achieved by reducing confounding factors from chronic health conditions which are more common in our population and attenuate the effects of individual-level socioeconomic and VA facility-level characteristics.

### Limitations

Our evaluation is focused on evaluating the association of area-level county-level SDH and COVID-19 test and test positivity of our unique Veteran population, who are on average are male, older, and have more comorbidities than the general US population, which limits generalizability.^13^ Furthermore, our evaluation does not assign weights to the county-level SDH relative to each other since there is no strong evidence to rigorously assign importance across categories.^1^ The association between COVID-19 infection risk and Veterans’ county-level SDH may be stronger than the estimated results presented here owing to the fact that some of the covariates adjusted for in this analysis may likely be mediators in the pathway, which would attenuate risk. Lastly, Veterans’ home address may not fully capture where Veterans spend most of their time which may result in exposure misclassification, however, we anticipate misclassification would be attenuated by county-level aggregation.

## Conclusion

In this evaluation of Veterans, we identified that county-level SDH factors influence COVID-19 infection risk, informing our understanding of how to improve care strategies, targeted interventions, policy, and resource allocation for Veterans.

## Data Availability

All data produced in the present work are contained in the manuscript

---

**Supplementary Table 1.**
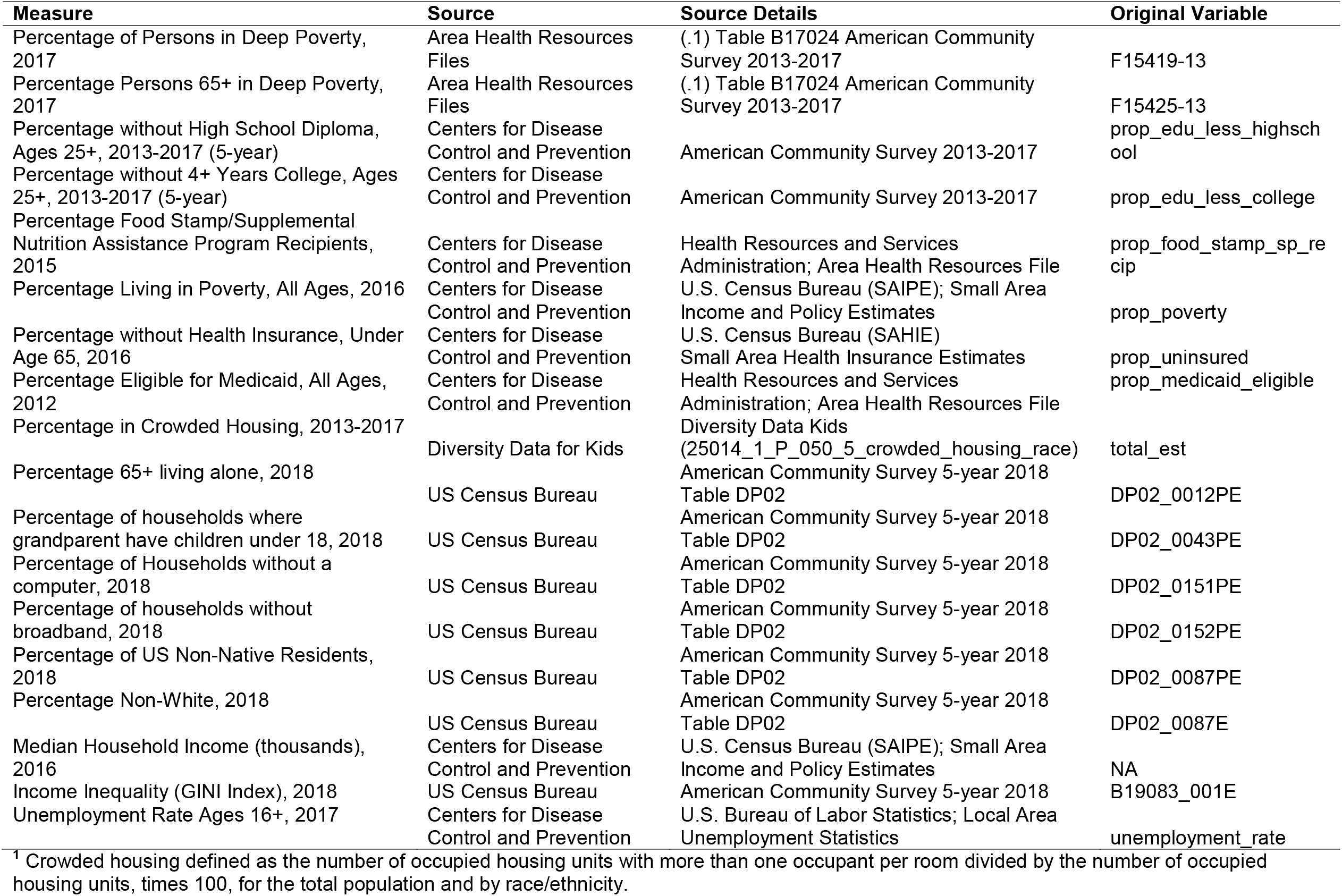
Data Dictionary for county-level socioeconomic measures.

| Measure | Source | Source Details | Original Variable |
| --- | --- | --- | --- |
| Percentage of Persons in Deep Poverty, 2017 | Area Health Resources Files | (.1) Table B17024 American Community Survey 2013-2017 | F15419-13 |
| Percentage Persons 65+ in Deep Poverty, 2017 | Area Health Resources Files | (.1) Table B17024 American Community Survey 2013-2017 | F15425-13 |
| Percentage without High School Diploma, Ages 25+, 2013-2017 (5-year) | Centers for Disease Control and Prevention | American Community Survey 2013-2017 | prop_edu_less_highschool |
| Percentage without 4+ Years College, Ages 25+, 2013-2017 (5-year) | Centers for Disease Control and Prevention | American Community Survey 2013-2017 | prop_edu_less_college |
| Percentage Food Stamp/Supplemental Nutrition Assistance Program Recipients, 2015 | Centers for Disease Control and Prevention | Health Resources and Services Administration; Area Health Resources File | prop_food_stamp_sp_recip |
| Percentage Living in Poverty, All Ages, 2016 | Centers for Disease Control and Prevention | U.S. Census Bureau (SAIPE); Small Area Income and Policy Estimates | prop_poverty |
| Percentage without Health Insurance, Under Age 65, 2016 | Centers for Disease Control and Prevention | U.S. Census Bureau (SAHIE) Small Area Health Insurance Estimates | prop_uninsured |
| Percentage Eligible for Medicaid, All Ages, 2012 | Centers for Disease Control and Prevention | Health Resources and Services Administration; Area Health Resources File | prop_medicaid_eligible |
| Percentage in Crowded Housing, 2013-2017 | Diversity Data for Kids | Diversity Data Kids (25014_1_P_050_5_crowded_housing_race) | total_est |
| Percentage 65+ living alone, 2018 | US Census Bureau | American Community Survey 5-year 2018 Table DP02 | DP02_0012PE |
| Percentage of households where grandparent have children under 18, 2018 | US Census Bureau | American Community Survey 5-year 2018 Table DP02 | DP02_0043PE |
| Percentage of Households without a computer, 2018 | US Census Bureau | American Community Survey 5-year 2018 Table DP02 | DP02_0151PE |
| Percentage of households without broadband, 2018 | US Census Bureau | American Community Survey 5-year 2018 Table DP02 | DP02_0152PE |
| Percentage of US Non-Native Residents, 2018 | US Census Bureau | American Community Survey 5-year 2018 Table DP02 | DP02_0087PE |
| Percentage Non-White, 2018 | US Census Bureau | American Community Survey 5-year 2018 Table DP02 | DP02_0087E |
| Median Household Income (thousands), 2016 | Centers for Disease Control and Prevention | U.S. Census Bureau (SAIPE); Small Area Income and Policy Estimates | NA |
| Income Inequality (GINI Index), 2018 | US Census Bureau | American Community Survey 5-year 2018 | B19083_001E |
| Unemployment Rate Ages 16+, 2017 | Centers for Disease Control and Prevention | U.S. Bureau of Labor Statistics; Local Area Unemployment Statistics | unemployment_rate |

<sup>1</sup> Crowded housing defined as the number of occupied housing units with more than one occupant per room divided by the number of occupied housing units, times 100, for the total population and by race/ethnicity.

